# Computational Phenotypes for Patients with Opioid-Related Disorders Presenting to the Emergency Department

**DOI:** 10.1101/2023.03.24.23287638

**Authors:** Richard Andrew Taylor, Aidan Gilson, Wade Schulz, Kevin Lopez, Patrick Young, Sameer Pandya, Andreas Coppi, David Chartash, David Fiellin, Gail D’Onofria

**Affiliations:** Section for Biomedical Informatics and Data Science, Yale University School of Medicine; Department of Emergency Medicine, Yale School of Medicine; Program of Computational Biology and Bioinformatics, Yale University; Department of Laboratory Medicine, Yale School of Medicine; Department of Internal Medicine, Yale School of Medicine; School of Medicine, University College Dublin - National University of Ireland, Dublin

**Keywords:** Opioid Use Disorder, Computational Phenotypes, Machine Learning, Opioid Pandemic

## Abstract

**Objective:** We aimed to discover computationally-derived phenotypes of opioid-related patient presentations to the emergency department (ED) via clinical notes and structured electronic health record (EHR) data.

**Methods:** This was a retrospective study of ED visits from 2013-2020 across ten sites within a regional healthcare network. We derived phenotypes from visits for patients 18 years of age with at least one prior or current documentation of an opioid-related diagnosis. Natural language processing was used to extract clinical entities from notes, which were combined with structured data within the EHR to create a set of features. We performed Latent Dirichlet allocation to identify topics within these features. Groups of patient presentations with similar attributes were identified by cluster analysis.

**Results:** In total 82,577 ED visits met inclusion criteria. The 30 topics discovered ranged from those related to substance use disorder, chronic conditions, mental health, and medical management. Clustering on these topics identified nine unique cohorts with one-year survivals ranging from 84.2-96.8%, rates of one-year ED returns from 9-34%, rates of one-year opioid event 10-17%, rates of medications for opioid use disorder from 17-43%, and a median Carlson comorbidity index of 2-8. Two cohorts of phenotypes were identified related to chronic substance use disorder, or acute overdose.

**Conclusions:** Our results indicate distinct phenotypic clusters with varying patient-oriented outcomes which provide future targets better allocation of resources and therapeutics. This highlights the heterogeneity of the overall population, and the need to develop targeted interventions for each population.

## Introduction

### Background

Opioid overdose deaths have risen to alarming levels, worsening dramatically during the COVID-19 pandemic.[1] For the first time, the CDC reported over 100,000 U.S. drug related deaths in a one-year period, a staggering 28.7% increase from the year prior, with over three-quarters of the deaths attributed to opioids.[2] With approximately 130 million patient visits per year and a federal mandate to ensure emergency care for anyone who presents regardless of their ability to pay, emergency departments (EDs) are an essential healthcare setting for reducing the morbidity and mortality associated with the overdose crisis.[3]

EDs serve as the primary healthcare venue for individuals following an opioid overdose, where patients discharged with a primary diagnosis of opioid overdose have a 1-year all-cause mortality rate of 5.5%.[4] Evidence-based interventions have offered a means to reduce the mortality rate, and are therefore crucial to implement. Despite the well-recognized importance of the ED in caring for patients with opioid-related illness, significant knowledge gaps persist concerning the characteristics and typology, or phenotypes, of opioid-related presentations to EDs and their associated outcomes.[5] Better understanding of visit presentations and their association with future events can enhance the interpretation of risk, help ED providers address gaps in treatment and care paradigms, and uncover areas for further research.

### Importance

Prior work on opioid related phenotypes in the ED has largely been confined to preconceived concepts (e.g., opioid use disorder (OUD), opioid overdose) using discrete structured data such as diagnostic codes or labs within electronic health records (EHRs).][5–7] While essential for trial cohort identification and quality measurement, these methods often fail to address more nuanced and latent representations of patients with opioid related conditions who often have complex, multifactorial disease processes and psychosocial interactions.[8, 9] Clinical notes within EHRs are an invaluable source of information about ED patients and visit presentations, often containing information not readily available in structured data fields (diagnostic codes, medication orders, etc.). Recent advances in natural language processing (NLP) concept extraction,[10] topic modeling,[11] and document level embedding[12] offer the opportunity to process notes in innovative ways and model the aforementioned unstructured information to enhance insight into patients with opioid conditions presenting to the ED. In turn, derived novel patient cohorts from these techniques may have differential associations with downstream care processes including treatment decisions, disposition decisions, utilization of healthcare resources (care coordination, etc.), and expenditure. Using NLP, we can better examine the characteristics and typology of patients presenting with opioid related conditions, making possible the derivation of novel treatment paradigms by phenotype.

### Goals of This Investigation

In this study, we aimed to discover phenotypes of ED patients with opioid conditions derived through NLP of ED clinical notes and structured data. We leveraged recent advances in NLP concept extraction to extract concepts from ED clinician and nursing notes and used Latent Dirichlet Allocation (LDA),[13] a topic modeling method, to develop visit level topics of patient presentations within ED clinical notes. With these phenotypes in hand, we used advanced clustering methods to identify unique cohorts within the population of patients with opioid conditions. Furthermore, examining the downstream outcomes of these cohorts we aimed to demonstrate their clinimetric potential within the ED.

## Methods

### Study Design and Setting

This was a retrospective observational cohort study of ED visits between 2013 and 2020 across ten sites within a regional healthcare network in the northeastern United States. These EDs include two non-academic urban, two academic urban, one pediatric urban, and five non-academic suburban sites. The sites encompass a geographic area of approximately 650 square miles and closely resemble the overall national population.[14] We included visits for all adult patients ≥ 18 years of age who had at least one opioid diagnosis (see Supplementary Table 1). This study followed STROBE reporting guidelines for observational studies.[15] Our institutional review board approved this study and waived the need for informed consent (HIC# 1602017249).

### Data Collection and Processing

Patient demographic and clinical data were extracted from the system-wide electronic health record (Epic, Verona, WI) using a centralized data warehouse (Helix). We initially identified a subset of ED notes representing patients with at least one current or prior opioid related diagnosis (the list of ICD10 codes used to identify patients is available in Supplement 1). Methodologic steps described below also necessitated the use of a representative subset of the entire ED population for use in the normalization of the opioid cohort concepts. For this step, an equal number of random ED encounters without an opioid diagnosis were identified along with associated ED notes.[16] Diagnoses were grouped according to the AHRQ Clinical Classification Software coding system.[17] Outpatient medications were mapped to the First DataBank Enhanced Therapeutic Classification System.[18]

### Clinical Concept Extraction

All subsequent steps of analysis are outlined in Figure 1. For clinical concept extraction, we use the Medical Concept Annotation Toolkit (MedCAT) that provides a novel self-supervised machine learning algorithm for extracting concepts to common data standards (UMLS, SNOMED-CT).[10] MedCAT overcomes many existing limitations noted during the comparison of tools such as MetaMap or cTAKES, including handling spelling mistakes and ambiguous concepts within a scalable solution.[19, 20] For the concept extraction task we use the full UMLS model trained on MIMIC III data. We further retrained this model on all 2.7 million ED physician notes and 6 million ED nursing notes written between 2013 and 2020 within the data warehouse. As the MIMIC II dataset is constructed from ICU encounters, this retraining allowed for the extraction of a broader set of clinical entities related to ED encounters and account for differences in documentation structure used in the MIMIC II dataset compared to our cohort. This model was then applied to both our opioid and random ED patient cohorts, and all text which was mapped to unique concept unique identifiers (CUIs) within each note were extracted. A knowledge source within the UMLS, the Semantic Network[21], was used to group all CUIs into larger categories (Semantic Types). We included all semantic type categories outlined in Supplemental Table 2. All other text in each note was removed. If a patient had multiple provider or nursing notes in a visit, all terms from each note were combined.

**Fig. 1.**
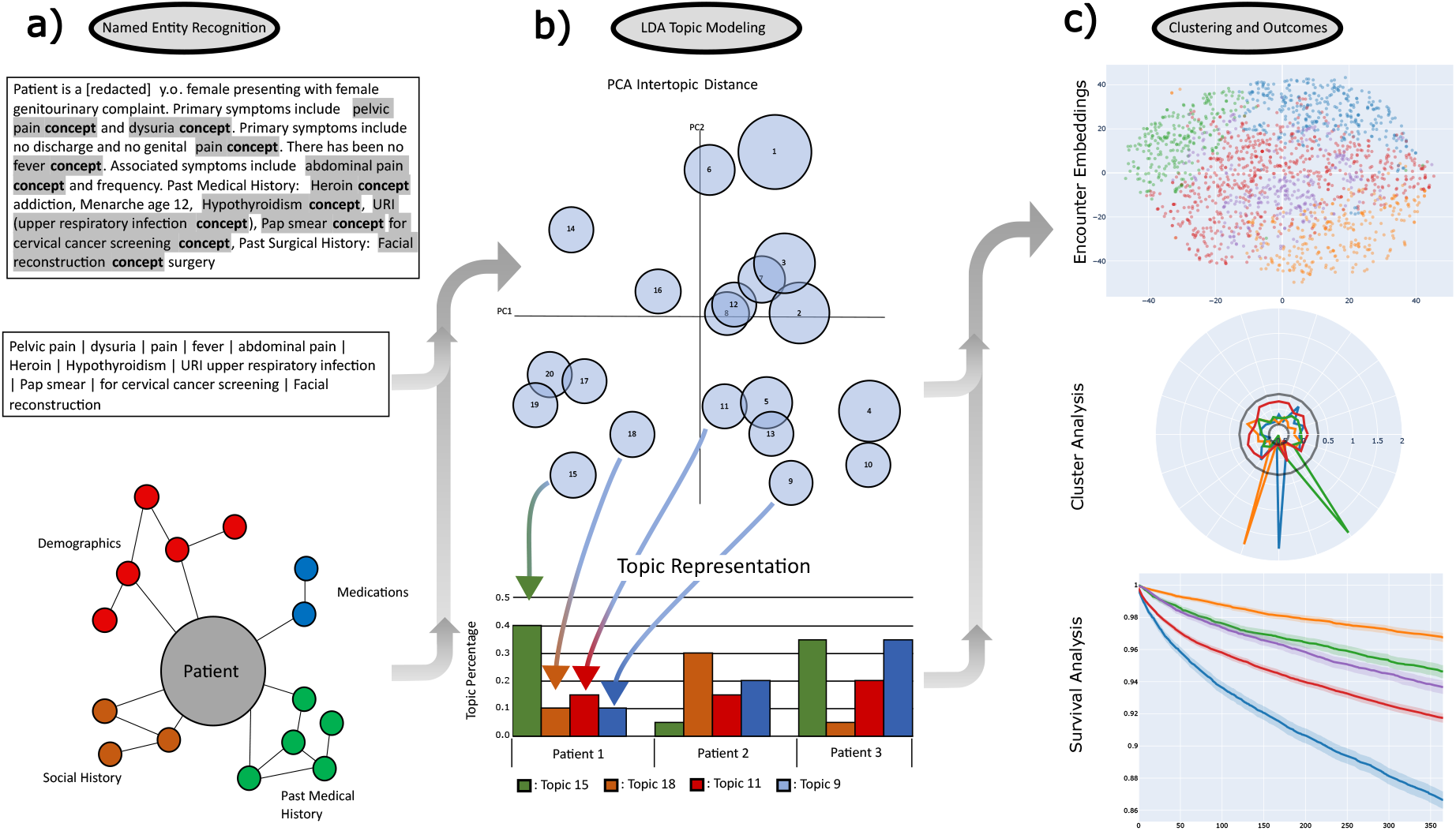
Workflow a) Entity extraction with clinical concept extraction using the trained MedCat model. Structured data labels combined with identified entities to complete the feature set. b) LDA Topic modeling of the extracted features converted to patient presentation embeddings. c) Clustering of topics, and outcomes analysis.

### Corpus Creation

Structured data was then added to the entity tags for each patient. The plaintext name of all structured information, for example “Low PaO2”, was added to each patient note if the code was present in the patient’s chart. For all continuous laboratory values, the plaintext name for the given laboratory value was added to the note along with the modifier “high” or “low” preceding the name if the patients value for the lab was greater than or less than two standard deviations from the mean of the matched random ED patient sample respectively.

### Topic Generation

We then used a Latent Dirichlet Allocation (LDA) model to embed the combined notes from the electronic health record (EHR) into latent feature space as implemented by the software package Mallet (2.0.8).[22] LDA is a generative statistical model used to collect individual discrete data elements of a corpus (e.g. words) into topics and facilitate the classification of collected data elements (e.g. documents).[13] We first identified and removed data elements (words) that appear in more than five percent of both the opioid exposed and random ED cohort datasets. This filtered out noise such as medical jargon related to common care processes and note templates. After removing these words, a contingency table was built from the remaining words to find keep words that have a p-value of less than 0.05 using a chi-square test for independence between cohorts. Keep words were then collected for each note within a patient encounter. Keep words from all notes within an encounter were then combined to create a term frequency corpus for each encounter. Mallet was used to generate the topics, each described by a set of keep words. The number of topics was chosen based on the model coherence plateau (see Supplementary Figure 1). For interpretability, topics were summarized with an overarching identifier through the analysis of keep words by expert consensus.[23] In Figure 2, word clouds were constructed for each topic with word size proportional to the probability of the data elements’ presence within that topic.

**Fig. 2.**
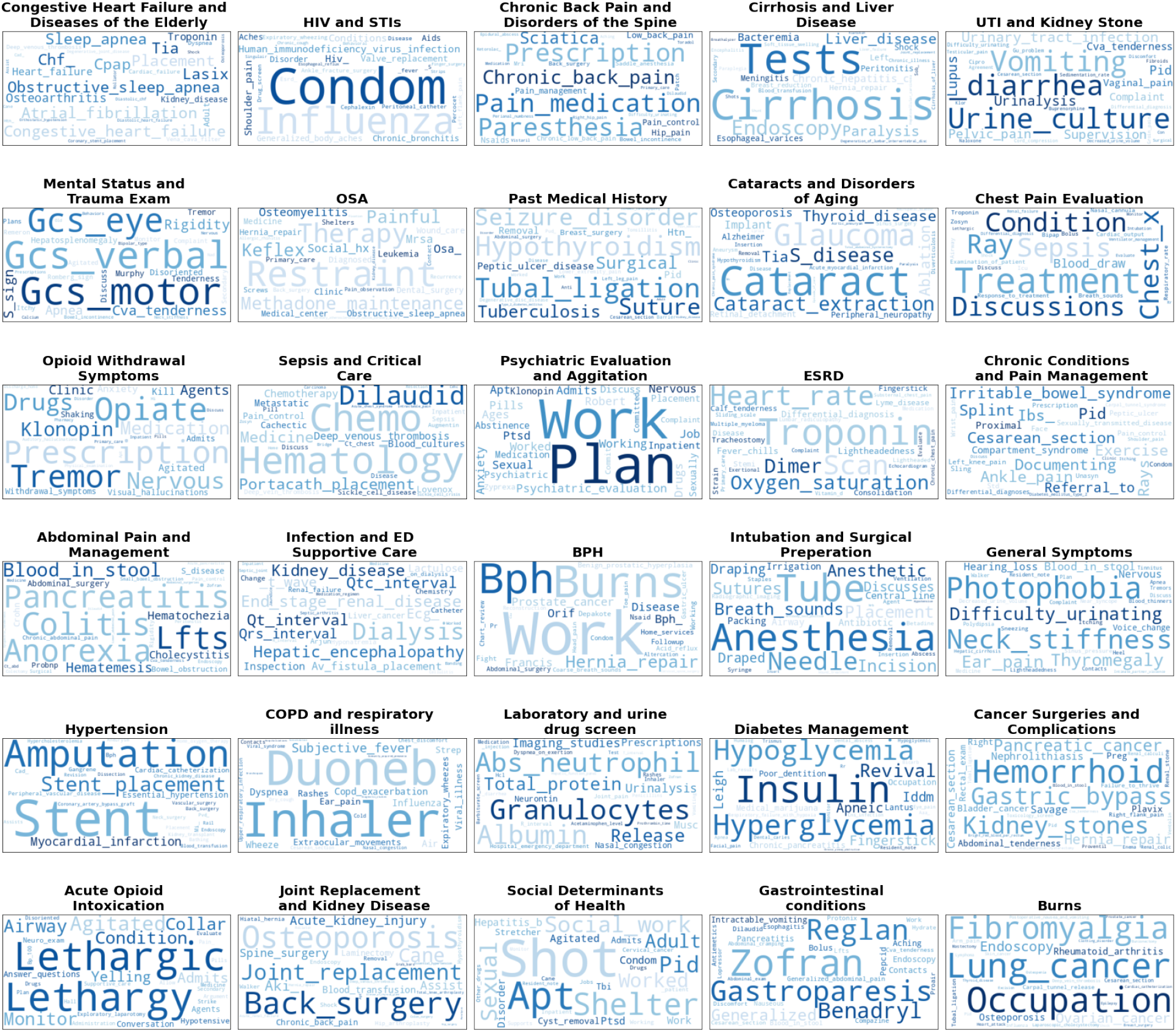
Wordclouds for the twenty topics identified by the LDA Mallet software. Word size is reflective of its importance within the topic. Topic titles were created by expert consensus to simplify discussion, but were not used in analysis.

We then used these topics to construct a n-dimensional embedding for each encounter within the opioid dataset, where n is the selected number of topics. The value of the ith dimension for a patient encounter is equal to the percentage that the ith topic is represented within the notes for that encounter, relative to all other topics. The sum of the embedding of all topics, and therefore all dimensions, will always equal one for a given patient encounter.

### Discovery and Validation of Opioid Phenotype Groups

Finally we used the topic embeddings of encounters to generate novel computational phenotypes using unsupervised learning. We used K-means clustering, to allow for the evaluation of cluster quality and assessed cluster cohesion, separation, and connectivity through silhouette coefficients, silhouette plots, and the Dunn metric respectively.[24] We analyzed the cluster performance through multiple external metrics: one-year survival following discharge, the average Charlson Comorbidity Index (CCI) of the cluster, 6-month and 12-month rate of all cause ED return, 6-month and 12-month rate of opioid related ED return, and the rates of outpatient prescription of naloxone, methadone and buprenorphine within each cluster.[25] K-means clustering was selected to allow for the evaluation of the clusters using these problem specific outcomes across a wide range of cluster number. Relative risk was used to qualify the presence of each structured variable within each cluster. For a cluster n and structured variable i, the relative rate (RR) was calculated using the formula:

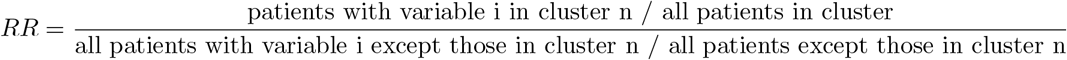

All analyses were conducted in Python (version 3.6) and R statistical software (version 3.4.3). The code employed in the analysis is accessible via the designated GitHub repository (https://cchteam.med.yale.edu/gitlab/computational-emergency-medicine/computational-based-phenotyping/-/tree/Publication_Branch). The data used in this study can be made available upon request.

## Results

In total, between March 1, 2013 and April 27, 2020, there were a total of 82,577 presentations of patients with prior or current documentation of an opioid related diagnosis which had associated structural related to ED presentation. All visit encounters contained a related provider note which was used in subsequent analysis regardless of presence of structured information. The characteristics of the patients with a history of opioid relative to the non-opioid cohort are shown in the Supplemental Table 3.

### Discovery and Validation of Opioid Phenotype Groups

LDA model coherence plateaued when twenty topics were selected. Figure 2 shows word clouds for each of the 30-topics identified, as well as the overarching identifier. The top 30 keepwords are shown, where word size is proportional to its importance in the topic. Topics, names here by expert consensus, ranged in areas from those directly related to substance use disorder (Opioid Withdrawal Symptoms, Acute Opioid Intoxication), past medical history (Congestive Heart Failure and DIseases of the Elderly, HIV and STIs, Chronic Back Pain and Disorders of the Spine, Cirrhosis and Liver Disease, etc.), mental health (Psychiatric Evaluation and Agitation), medical process (Laboratory and urine drug screen, Mental Status and Trauma exam), among others.

### Clustering

Figure 3 shows clustering of the embeddings for each patient encounter formed from the representation of each LDA topic within the patient encounter. When optimized for each clustering metric, K-Means clustering identified 9 unique clusters. Cluster 1 (blue) contained patients defined by the topic Laboratory and urine drug screen. Cluster 2 (orange) was defined by mixed GI conditions and abdominal pain. The remaining clusters were 3 (green) mental status and trauma evaluation, 4 (red) mixed representation across all topics, 5 (purple) psychiatric evaluation, 6 (brown) join replacement and kidney disease, 7 (pink) acute opioid intoxication, 8 (gray) general symptoms, 9 (yellow) social determinants of health.

**Fig. 3.**
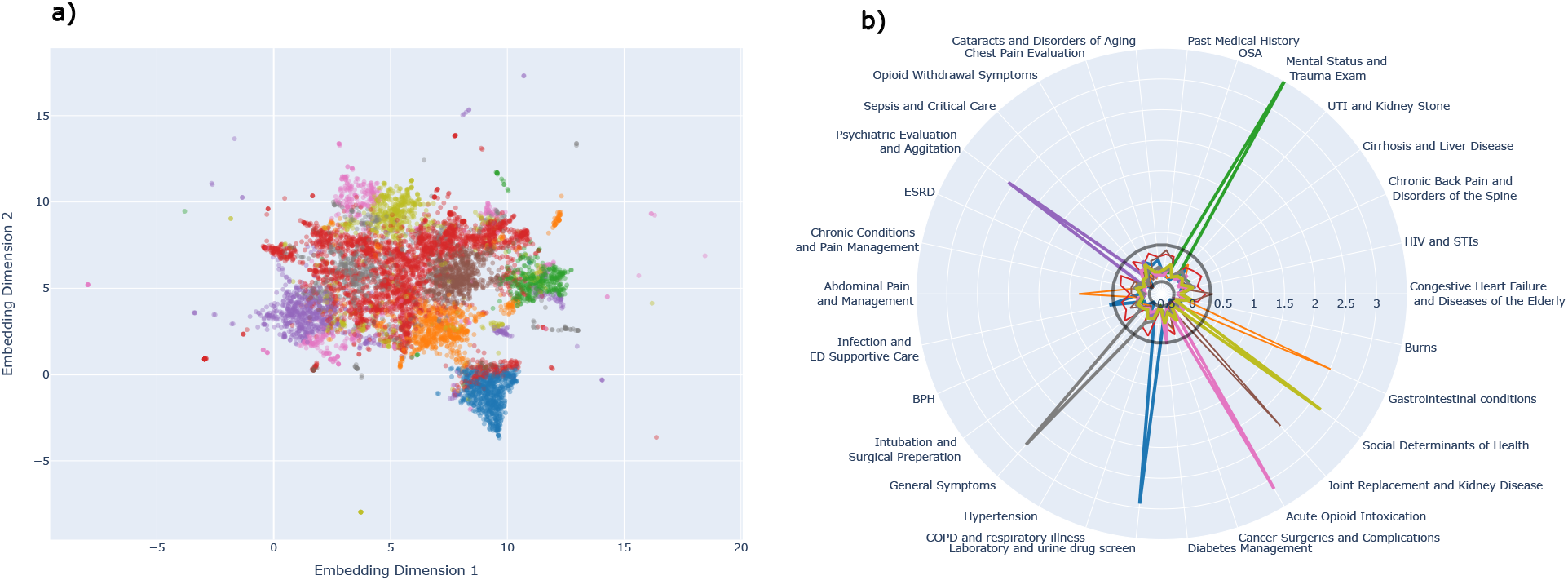
(left) 2-D K-Means and Agglomerative clustering of LDA topic embeddings with tSNE projection. (right) Representation of the LDA topics within each of the clusters using absolute mean standardized deviation of each topic relative to the overall cohort of opioid patients for the structured data components.

Table 1 identifies the demographic data for each cluster as well as the rates of mental health disorders, substance use disorder, and sequelae of IV drug use. Figure 4 similarly shows the relative risk each cluster has for the structured data components, as well as the relative risk for different chief complaints. Figure 5 shows outcome metrics which can further separate clusters. Cluster 1 is a more white cohort with lower rates of mental health disorders. Cluster 2 is a female population with a high Emergency Severity Index (ESI). Cluster 3 is a male population with increased Medicaid usage. Cluster 4 is a slightly more African American population. Cluster 5 is younger male population with increased Medicaid usage and high rates of suicide and substance use disorder. Cluster 6 is an older, female cohort with high Medicare usage and high rates of admission as well as a high average CCI. Cluster 7 is a younger heavily male population with emergent ED presentations. Cluster 8 is a younger male population with decreased naloxone prescription. Finally, cluster 9 is equally male and female with very high rates of alcohol and other substance use disorder. Final cluster descriptions are shown in Table 2.

**Table 1.**
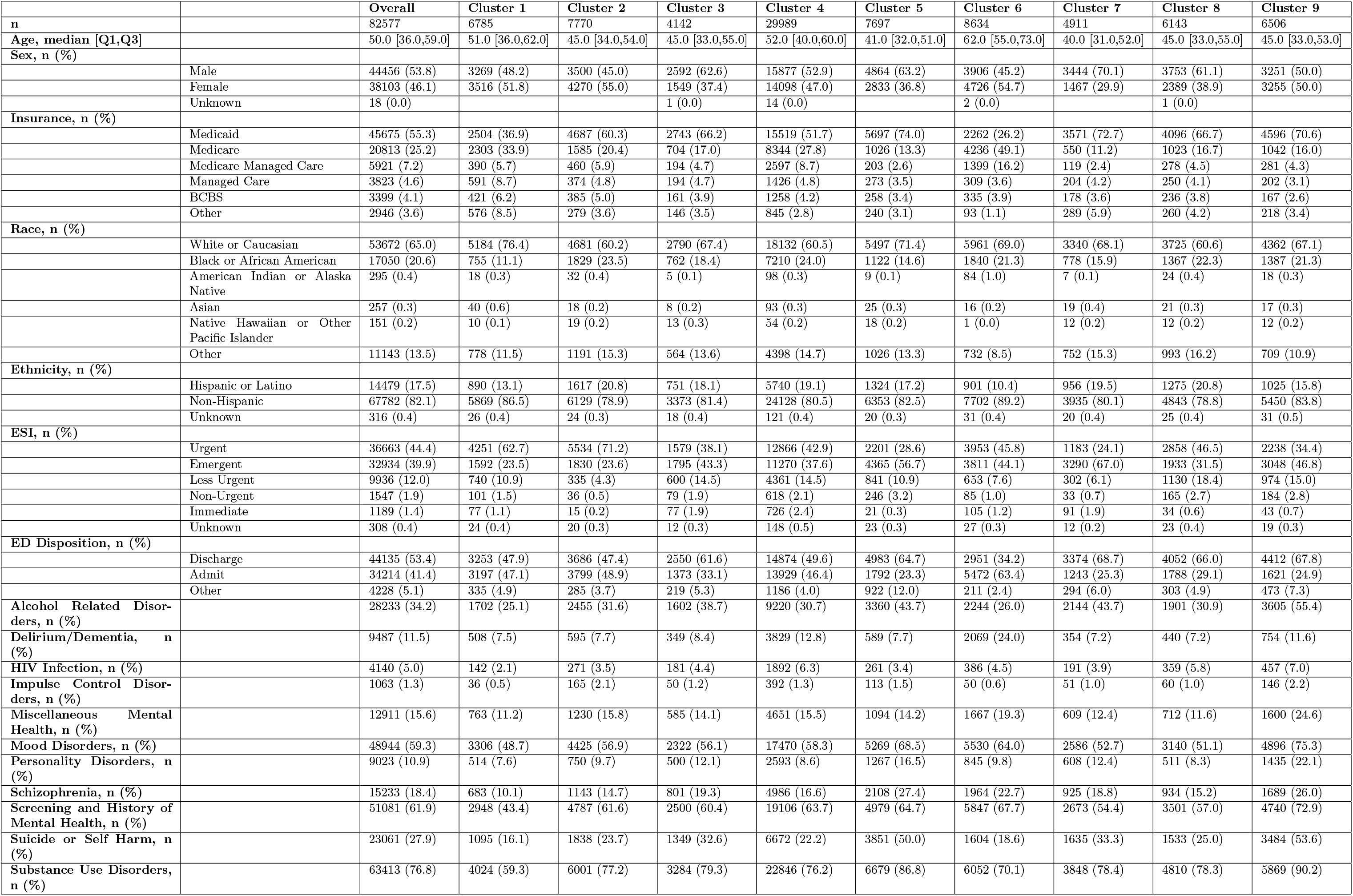
Feature Breakdown of Characteristics for Clusters Identified by K-Means

**Table 2.** Cluster Descriptions

| Cluster | Description |
| --- | --- |
| 1 (blue) | White female population with increased Medicare use, lower rates of mental health disorders and substance use disorder, and moderate ED acuity. |
| 2 (orange) | Younger female population with GI complaints, nausea and vomiting on presentation to ED, with a high ESI. |
| 3 (green) | Male population with high Medicaid usage and trauma evaluation. |
| 4 (red) | Black population with higher comorbidity burden, immediate ESI, high rates of HIV, presenting with constitutional complaints (shortness of breath, back pain, leg pain, leg swelling, fever, and cough). |
| 5 (purple) | Young male population with high substance use disorder, and suicide rates, presenting as suicidal or in need of a psychiatric evaluation. |
| 6 (brown) | Older female population with significant Medicare usage, high rates of dementia, a significant comorbidity burden, presenting with altered mental status, fall and/or weakness. |
| 7 (pink) | Young male population with emergent presentation, high rates of ED discharge, presenting with acute intoxication or overdose. |
| 8 (gray) | Younger male non-white population with low urgency ESI, presenting with altered mental status, dizziness, and/or headache. |
| 9 (yellow) | A population with high rates of teenagers, with significant substance use disorder and mental health disorders and factors related to social determinants of health. |

**Fig. 4.**
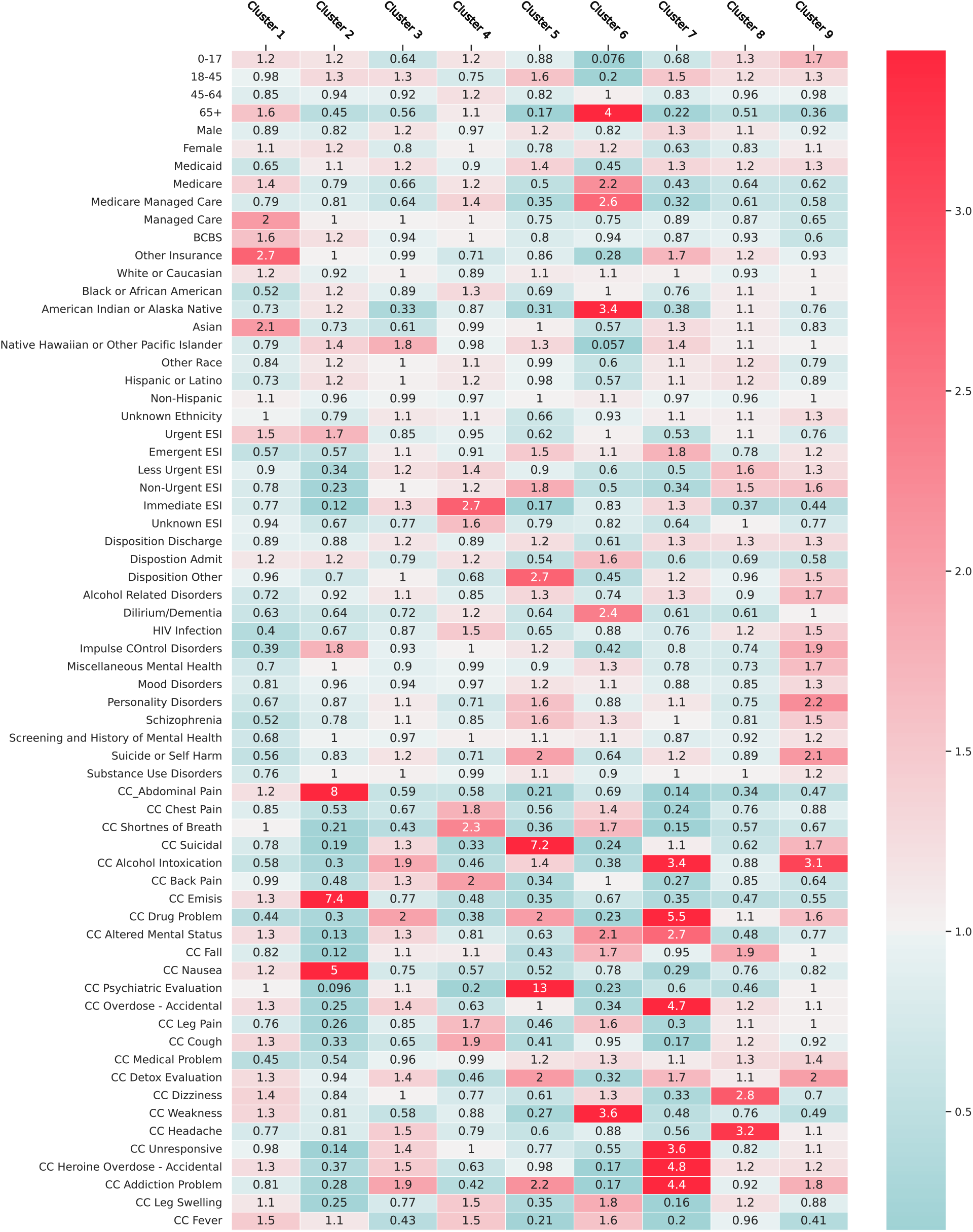
Heatmap of the relative risk of the structured data component for each cluster. Relative to the entire population of opioid patients, red cells mark increased relative risk, while teal cells mark decreased relative risk. Each column therefore represents the information with a cluster that differs most significantly from the overall OUD population.

**Fig. 5.**
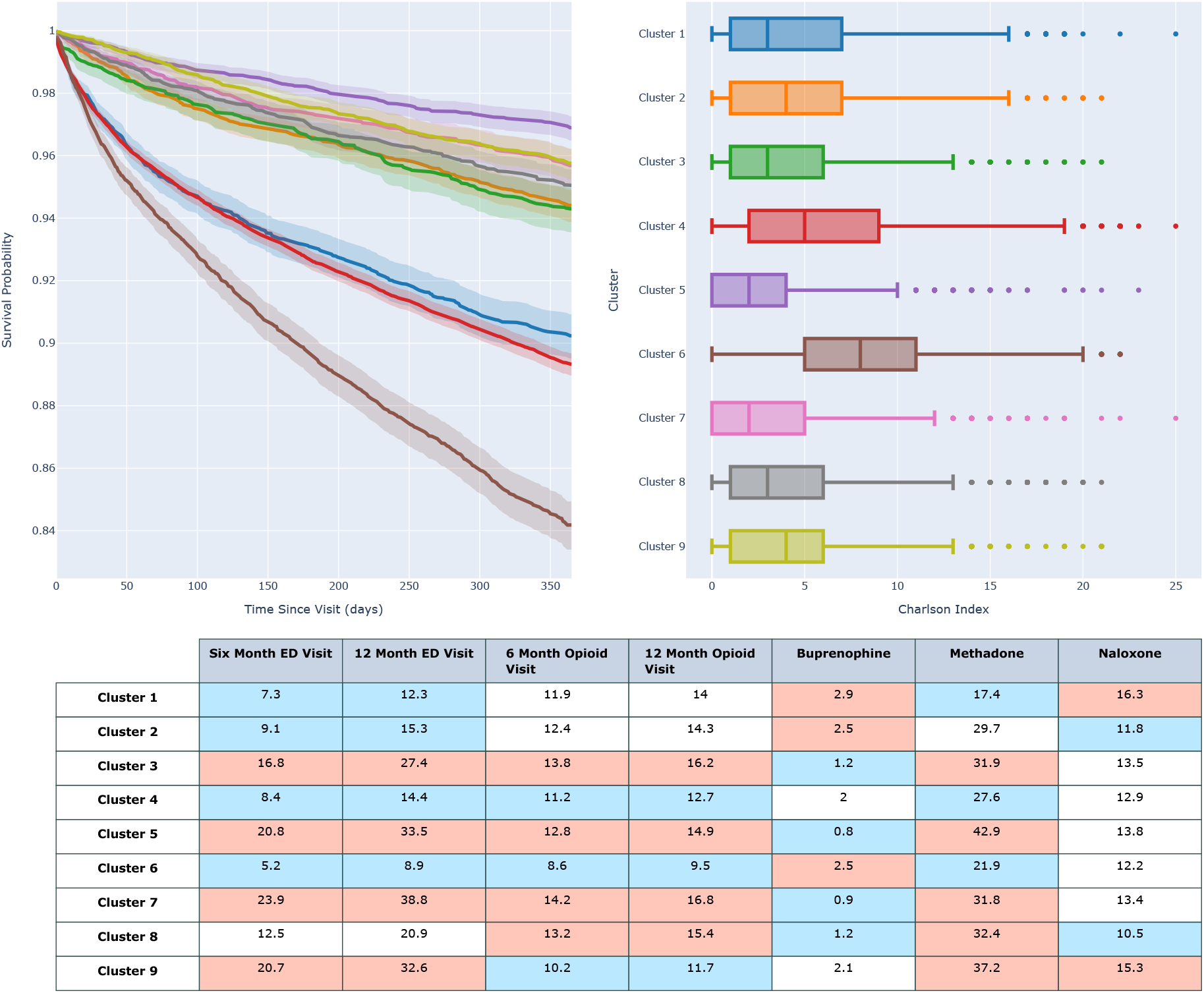
a) Kaplan-Meier curve showing two-year survival following hospital discharge. b) Box and Whisker plot of the distribution of the CCI score for the patients within each cohort. c) Rates of 6 and 12 month ED return for all causes, rates of 6 and 12 month opioid related event, rates of prescription of Beuprenorphine, Methadone, and Naloxone. Relative to the entire dataset, red cells have statistically increased rates by chi-square test vs the remaining population, blue cells have statistically decreased rates by the same metric.

### Outcomes Analysis

Figure 5 displays the results of the outcomes analysis. Cluster 5 had a significantly higher one-year survival (96.9%) and lower median CCI (2) than all other clusters (p¡0.05). Clusters 2, 3, 7, 8, and 9 had overlapping survival functions and were not separable (94.3%-95.7%). Clusters 1 and 4 had lower survival rates (90.2% and 89.3%), although cluster 4 has a significantly higher median CCI (5 vs 3). Finally, Cluster 6 has the lowest survival rate (84.2%) and highest median CCI (8).

Clusters 3,5,7, and 9 have the highest rates of all cause ED return in both the 6- and 12-month windows, however all clusters, with the exception of Cluster 6 have similar rates of ED return related to an opioid condition. Cluster 5 has the highest rates of outpatient methadone usage, (42.9%). Cluster 6 is the only cluster with similar rates of methadone and naloxone prescription (17.4% and 16.3% respectively), where no other cluster has naloxone prescriptions exceeding 60% of methadone prescriptions.

## Discussion

The opioid epidemic presents a severe and multifaceted challenge that impacts patients in various ways, resulting in a considerable number of opioid-related fatalities, non-fatal overdoses, and secondary consequences.[26, 27] A crucial approach to addressing this issue involves identifying the diverse patient populations affected by the opioid epidemic and their unique characteristics.[5] In this study, we aimed to uncover these populations by developing opioid-related ED phenotypes through the integration of structured patient information in the EHR, unstructured data contained in notes, and cluster analysis. Our findings reveal nine distinct opioid-related cohorts, each characterized by unique profiles of demographic, diagnostic, pharmaceutical, and healthcare utilization. These cohorts exhibit larger trends, such as three younger male populations with high acute severity and emergency department (ED) usage (Clusters 3, 5, and 7), two populations with significant comorbidity burden (Clusters 4 and 6), two predominantly female populations (Clusters 3 and 6), and two populations with notable substance use disorder (Clusters 5 and 9). This granular classification of patients facilitates targeted interventions for specific groups and offers a hierarchical approach for patient classification.

Previous studies on the phenotyping of ED patients presenting for opioid-related reasons primarily focused on differentiating opioid use disorder (OUD) from opioid overdose.[6, 7] The primary objective of these investigations centered on the precise identification of patient cohorts within Electronic Health Records (EHR) for the purposes of subject selection and pragmatic clinical trials. Methodologically, however, these studies faced limitations in their capacity to further discriminate between distinct subgroups. A few other studies have explored broader, computationally driven phenotypes of opioid related conditions. In 2019 Afshar et al. described four subgroups of opioid use disorder, patients with known disorder, patients with low socioeconomic status and psychosis, patients with alcohol use disorder, and patients with incidental opioid misuse.[28] Additionally, Schirle et al developed both a comorbidity and a NLP based scoring system for identifying the risk of OUD from EHR data.[29] These models identified structured elements with a correlation for OUD however they were not synthesized into complete phenotypes.

Our approach and findings diverge from those presented in previous literature by undertaking a more comprehensive search for opioid-related phenotypes focused on emergency department (ED) presentations. We accomplish this by employing a combination of structured and unstructured data sources, thereby capturing a more diverse and representative sample. Additionally, we extend our analysis by associating these phenotypes with crucial clinical and therapeutic outcomes, enabling an understanding of the relationship between opioid-related conditions and their respective consequences.

We identified four distinct groups within the chronic use disorder cohort (clusters 1, 4, 6, and 9), each exhibiting unique outcomes, characteristics, and prescribing patterns. For instance, Cluster 6, the older female cohort, has a one-year survival rate 4% lower than the next phenotype within the chronic opioid use category (Cluster 4) and a significantly high comorbidity burden as measured by the average Charlson Comorbidity Index (CCI). However, this cluster also demonstrates the lowest ED return rates within the timeframes examined. Our analysis also identified three cohorts (Clusters 3, 5, and 7) more significantly associated with acute opioid overdose, with demographic breakdowns resembling those described in previous literature.[30] These cohorts exhibit the highest ED return rates among all identified clusters but also some of the lowest one-year mortalities, likely due to the lower average age compared to other cohorts.

The identification of these subgroups and their unique outcomes underscores the necessity of developing targeted interventions for each population. The substantial burden that the opioid epidemic has placed on both the general population and the healthcare system makes it challenging to provide patient-specific interventions when providers are overwhelmed with the number and variety of patients they encounter. The phenotypes discovered in this study can offer an initial framework for physicians to categorize patients under their care, leading to better resource allocation for patients with the highest mortality (Cluster 6), targeted integration of patients with the highest ED utilization with outpatient follow-up (Clusters 3, 5, and 7), distribution of naloxone and other life-saving medications to patients most at risk of acute overdose (Clusters 3, 5, and 7), and assessment of a patient’s comorbidities at the initial prescription of an opioid to evaluate the risk of abuse related to underlying pain-causing conditions (Clusters 2 and 6).

In addition to the primary findings, the utilization of topic modeling techniques on provider notes has corroborated numerous intriguing opioid-related topics that span a diverse range of areas. This validation of previously established relationships further emphasizes the complex interplay between opioid use and various health conditions, strengthening the evidence base for these associations and reinforcing their significance in the context of opioid-related research and patient care. These areas include social determinants of health, psychiatric evaluations, and the management of chronic conditions such as diabetes, congestive heart failure, and chronic obstructive pulmonary disease.[31–35] Furthermore, several identified topics demonstrate a connection to pain-related conditions, including kidney stones, orthopedic procedures, surgeries, and burns. These discovered associations offer a deeper understanding of the intricate relationship between opioid use and various health conditions. For example, the association between diabetes management and opioid use may expose potential challenges in medication adherence and blood sugar regulation for patients dealing with simultaneous opioid dependencies. Likewise, the connection between chronic pain and opioid use underscores the necessity for alternative pain management strategies, emphasizing the importance of addressing the underlying causes of pain while minimizing opioid dependency.[36] The association between infections and opioid use further accentuates the well-documented susceptibility to infectious complications, particularly among intravenous drug users.[37] This relationship underscores the importance of vigilant monitoring and the implementation of preventive measures to mitigate the risk of infections in this vulnerable population. Finally, the association with opioid withdrawal symptoms highlights the demand for efficacious management strategies to mitigate the discomfort and obstacles encountered by patients during the withdrawal process. The phenotypes identified in this study hold significant potential for a variety of applications and future use in both clinical and research settings. By providing a more granular classification of patients affected by the opioid epidemic, these phenotypes enable healthcare providers to tailor interventions and treatment strategies based on the unique characteristics and needs of each subgroup. This targeted approach can lead to more effective resource allocation, improved patient outcomes, and a better understanding of the underlying factors contributing to opioid-related conditions. Furthermore, the phenotypes serve as a valuable resource for future research, aiding in the development and evaluation of new therapeutic approaches, preventive measures, and public health initiatives. As our understanding of the multifaceted nature of opioid-related conditions continues to grow, the phenotypes can be refined and expanded upon to better capture the evolving landscape of the opioid epidemic, ultimately contributing to a more comprehensive approach to addressing this complex public health challenge.

### Limitations

This study has several limitations. First, there is an inherent trade-off between specificity and interpretability of the identified phenotypes. Our topics represent broad level phenotypes of patients, but not all patients fall into one of these categories, as evidenced by cluster 4, which is not well defined. Increased specificity of the phenotypes, which may also necessitate an increased number, would allow for more precise stratification of patient outcomes, but would complicate grouping new patients into their phenotypes as well as complicate the ease of assigning new patients to a phenotype. As the assignment of phenotypes to a novel patient encounter is mutually exclusive, increased specificity of the phenotypes may result in the oversimplification of an encounter to a single phenotype when it shares elements from multiple. Second, there is a chance for the injection of preconceived biases when identifying LDA topics.[23] Although we found topic coherence plateaued at 30 topics, similar coherence levels were achieved at lower topic number and through variation of other hyperparameters inherent in the algorithm. Final selection of topics was based on clinical judgment of topic coherence, which may be susceptible to biases in expected topics. We have included the topic descriptions for other topic numbers in the supplementary data. Third, the concept extraction technique used is limited by the ability of the natural language processing (NLP) algorithms to accurately identify and extract relevant information from the clinical notes. This may result in important information being overlooked or misclassified. Fourth, the use of Latent Dirichlet Allocation (LDA) for topic modeling is limited by the assumption that each document can be represented as a mixture of topics and that the topics themselves are generated from a Dirichlet distribution.[38] This may not accurately capture the complexity of clinical notes and EHR data and the actual distribution of topics within the encounters may not be completely represented by a Dirichlet distribution. Fifth, the limitations of labeling theLDA topics can lead to inaccuracies in the interpretation of the phenotypes. This may result from subjective judgments and the difficulty in accurately categorizing complex information into discrete concepts. Sixth, the limitations of unsupervised clustering and the optimization of clustering size can impact the validity of the phenotype groups identified. This includes issues such as the sensitivity of the clustering algorithm to the choice of initial conditions and the difficulty in determining the optimal number of clusters. Finally, the study was conducted within a single healthcare network in the northeastern United States, which may limit the ability to generalize the results to other populations or healthcare systems.

## Conclusions

Our results indicate distinct phenotypic clusters with varying patient-oriented outcomes providing future targets better allocation of resources and therapeutics. Not only does this analysis separate the patients into acute and chronic populations, but also provides further stratification within each of these cohorts. Although all patients share the burden of opioid related conditions, our analysis shows that past this similarity there is significant heterogeneity within the population. This separability of phenotypes demonstrates the need for specific evidence-based interventions for each phenotype. The identified subgroups provide a starting point for physicians to organize and better understand the patients under their care. Future work is needed to determine the most effective interventions for each population.

## Data Availability

All data produced in the present study are available upon reasonable request to the authors

