## Supplements for "Computational Phenotypes for Patients with Opioid-Related Disorders Presenting to the Emergency Department"

### Supplementary Tables:

Supplementary Table 1: ICD10 Codes used to Identify Patients with ED Opioid Exposure.

| ICD-10-CM Code | ICD-10-CM Description |
|---|---|
| F11.10 | Opioid abuse, uncomplicated |
| F11.120 | Opioid abuse with intoxication, uncomplicated |
| F11.121 | Opioid abuse with intoxication delirium |
| F11.122 | Opioid abuse with intoxication with perceptual disturbance |
| F11.129 | Opioid abuse with intoxication, unspecified |
| F11.14 | Opioid abuse with opioid-induced mood disorder |
| F11.150 | Opioid abuse with opioid-induced psychotic disorder with delusions |
| F11.151 | Opioid abuse with opioid-induced psychotic disorder with hallucinations |
| F11.159 | Opioid abuse with opioid-induced psychotic disorder, unspecified |
| F11.181 | Opioid abuse with opioid-induced sexual dysfunction |
| F11.182 | Opioid abuse with opioid-induced sleep disorder |
| F11.188 | Opioid abuse with other opioid-induced disorder |
| F11.19 | Opioid abuse with unspecified opioid-induced disorder |
| F11.20 | Opioid dependence, uncomplicated |
| F11.220 | Opioid dependence with intoxication, uncomplicated |
| F11.221 | Opioid dependence with intoxication delirium |
| F11.222 | Opioid dependence with intoxication with perceptual disturbance |
| F11.229 | Opioid dependence with intoxication, unspecified |
| F11.23 | Opioid dependence with withdrawal |
| F11.24 | Opioid dependence with opioid-induced mood disorder |

| | |
|---|---|
| F11.250 | Opioid dependence with opioid-induced psychotic disorder with delusions |
| F11.251 | Opioid dependence with opioid-induced psychotic disorder with hallucinations |
| F11.259 | Opioid dependence with opioid-induced psychotic disorder, unspecified |
| F11.281 | Opioid dependence with opioid-induced sexual dysfunction |
| F11.282 | Opioid dependence with opioid-induced sleep disorder |
| F11.288 | Opioid dependence with other opioid-induced disorder |
| F11.29 | Opioid dependence with unspecified opioid-induced disorder |
| F11.90 | Opioid use, unspecified, uncomplicated |
| F11.920 | Opioid use, unspecified, with intoxication, uncomplicated |
| F11.921 | Opioid use, unspecified, with intoxication delirium |
| F11.922 | Opioid use, unspecified, with intoxication with perceptual disturbance |
| F11.929 | Opioid use, unspecified, with intoxication, unspecified |
| F11.93 | Opioid use, unspecified with withdrawal |
| F11.94 | Opioid use, unspecified with opioid-induced mood disorder |
| F11.950 | Opioid use, unspecified with opioid-induced psychotic disorder with delusions |
| F11.951 | Opioid use, unspecified with opioid-induced psychotic disorder with hallucinations |
| F11.959 | Opioid use, unspecified with opioid-induced psychotic disorder, unspecified |
| F11.981 | Opioid use, unspecified with opioid-induced sexual dysfunction |
| F11.982 | Opioid use, unspecified with opioid-induced sleep disorder |
| F11.988 | Opioid use, unspecified with other opioid-induced |
| F11.99 | Opioid use, unspecified with unspecified opioid-induced disorder |
| T40.0X1A | Poisoning by opium, accidental (unintentional), initial encounter |
| T40.0X1D | Poisoning by opium, accidental (unintentional), subsequent encounter |
| T40.0X1S | Poisoning by opium, accidental (unintentional), sequela |
| T40.0X4A | Poisoning by opium, undetermined, initial encounter |
| T40.0X4D | Poisoning by opium, undetermined, subsequent encounter |

| | |
|---|---|
| T40.0X4S | Poisoning by opium, undetermined, sequela |
| T40.0X5A | Adverse effect of opium, initial encounter |
| T40.0X5D | Adverse effect of opium, subsequent encounter |
| T40.0X5S | Adverse effect of opium, sequela |
| T40.1X1A | Poisoning by heroin, accidental (unintentional), initial encounter |
| T40.1X1D | Poisoning by heroin, accidental (unintentional), subsequent encounter |
| T40.1X1S | Poisoning by heroin, accidental (unintentional), sequela |
| T40.1X4A | Poisoning by heroin, undetermined, initial encounter |
| T40.1X4D | Poisoning by heroin, undetermined, subsequent encounter |
| T40.1X4S | Poisoning by heroin, undetermined, sequela |
| T40.2X1A | Poisoning by other opioids, accidental (unintentional), initial encounter |
| T40.2X1D | Poisoning by other opioids, accidental (unintentional), subsequent encounter |
| T40.2X1S | Poisoning by other opioids, accidental (unintentional), sequela |
| T40.2X4A | Poisoning by other opioids, undetermined, initial encounter |
| T40.2X4D | Poisoning by other opioids, undetermined, subsequent encounter |
| T40.2X4S | Poisoning by other opioids, undetermined, sequela |
| T40.2X5A | Adverse effect of other opioids, initial encounter |
| T40.2X5D | Adverse effect of other opioids, subsequent encounter |
| T40.2X5S | Adverse effect of other opioids, sequela |
| T40.3X1A | Poisoning by methadone, accidental (unintentional), initial encounter |
| T40.3X1D | Poisoning by methadone, accidental (unintentional), subsequent encounter |
| T40.3X1S | Poisoning by methadone, accidental (unintentional), sequela |
| T40.3X4A | Poisoning by methadone, undetermined, initial encounter |
| T40.3X4D | Poisoning by methadone, undetermined, subsequent encounter |
| T40.3X4S | Poisoning by methadone, undetermined, sequela |
| T40.3X5A | Adverse effect of methadone, initial encounter |

| | |
|---|---|
| T40.3X5D | Adverse effect of methadone, subsequent encounter |
| T40.3X5S | Adverse effect of methadone, sequela |
| T40.4X1A | Poisoning by synthetic narcotics, accidental (unintentional), initial encounter |
| T40.4X1D | Poisoning by synthetic narcotics, accidental (unintentional), subsequent encounter |
| T40.4X1S | Poisoning by synthetic narcotics, accidental (unintentional), sequela |
| T40.4X4A | Poisoning by synthetic narcotics, undetermined, initial encounter |
| T40.4X4D | Poisoning by synthetic narcotics, undetermined, subsequent encounter |
| T40.4X4S | Poisoning by synthetic narcotics, undetermined, sequela |
| T40.4X5A | Adverse effect of synthetic narcotics, initial encounter |
| T40.4X5D | Adverse effect of synthetic narcotic, subsequent encounter |
| T40.4X5S | Adverse effect of synthetic narcotic, sequela |
| T40.601A | Poisoning by unspecified narcotics, accidental (unintentional), initial encounter |
| T40.601D | Poisoning by unspecified narcotics, accidental (unintentional), subsequent encounter |
| T40.601S | Poisoning by unspecified narcotics, accidental (unintentional), sequela |
| T40.604A | Poisoning by unspecified narcotics, undetermined, initial encounter |
| T40.604D | Poisoning by unspecified narcotics, undetermined, subsequent encounter |
| T40.604S | Poisoning by unspecified narcotics, undetermined, sequela |
| T40.605A | Adverse effect of unspecified narcotics, initial encounter |
| T40.605D | Adverse effect of unspecified narcotics, subsequent encounter |
| T40.605S | Adverse effect of unspecified narcotics, sequela |
| T40.691A | Poisoning by other narcotics, accidental (unintentional), initial encounter |
| T40.691D | Poisoning by other narcotics, accidental (unintentional), subsequent encounter |
| T40.691S | Poisoning by other narcotics, accidental (unintentional), sequela |
| T40.694A | Poisoning by other narcotics, undetermined, initial encounter |
| T40.694D | Poisoning by other narcotics, undetermined, subsequent encounter |
| T40.694S | Poisoning by other narcotics, undetermined, sequela |

| | |
|---|---|
| T40.695A | Adverse effect of other narcotics, initial encounter |
| T40.695D | Adverse effect of other narcotics, subsequent encounter |
| T40.695S | Adverse effect of other narcotics, sequela |
| T40.0X2A | Poisoning by opium, intentional self-harm, initial encounter |
| T40.0X2D | Poisoning by opium, intentional self-harm, subsequent encounter |
| T40.0X2S | Poisoning by opium, intentional l self-harm, sequela |
| T40.0X3A | Poisoning by opium, assault, initial encounter |
| T40.0X3D | Poisoning by opium, assault subsequent encounter |
| T40.0X3S | Poisoning by opium, , assault, sequela |
| T40.1X2A | Poisoning by heroin, intentional self-harm, initial encounter |
| T40.1X2D | Poisoning by heroin, intentional self-harm, subsequent encounter |
| T40.1X2S | Poisoning by heroin, intentional self-harm, sequela |
| T40.1X3A | Poisoning by heroin, assault, initial encounter |
| T40.1X3D | Poisoning by heroin, assault, subsequent encounter |
| T40.1X3S | Poisoning by heroin, assault, sequela |
| T40.2X2A | Poisoning by other opioids, intentional self-harm, initial encounter |
| T40.2X2D | Poisoning by other opioids, intentional self-harm, subsequent encounter |
| T40.2X2S | Poisoning by other opioids, intentional self-harm, sequela |
| T40.2X3A | Poisoning by other opioids, assault, initial encounter |
| T40.2X3D | Poisoning by other opioids, assault, subsequent encounter |
| T40.2X3S | Poisoning by other opioids, assault, sequela |
| T40.3X2A | Poisoning by methadone, intentional self-harm, initial encounter |
| T40.3X2D | Poisoning by methadone, intentional self-harm, subsequent encounter |
| T40.3X2S | Poisoning by methadone, intentional self-harm, sequela encounter |
| T40.3X3A | Poisoning by methadone, assault, initial encounter |
| T40.3X3D | Poisoning by methadone, assault, subsequent encounter |

| | |
|---|---|
| T40.3X3S | Poisoning by methadone, assault, sequela encounter |
| T40.4X2A | Poisoning by other synthetic narcotics, intentional self-harm, initial encounter |
| T40.4X2D | Poisoning by other synthetic narcotics, intentional self-harm, subsequent encounter |
| T40.4X2S | Poisoning by other synthetic narcotics, intentional self-harm, sequela |
| T40.4X3A | Poisoning by other synthetic narcotics, assault, initial encounter |
| T40.4X3D | Poisoning by other synthetic narcotics, assault, subsequent encounter |
| T40.4X3S | Poisoning by other synthetic narcotics, assault, sequela |
| T40.602A | Poisoning by unspecified narcotics, intentional self-harm, initial encounter |
| T40.602D | Poisoning by unspecified narcotics, intentional self-harm, subsequent encounter |
| T40.602S | Poisoning by unspecified narcotics, intentional self-harm, sequela encounter |
| T40.603A | Poisoning by unspecified narcotics, assault, initial encounter |
| T40.603D | Poisoning by unspecified narcotics, assault, subsequent encounter |
| T40.603S | Poisoning by unspecified narcotics, assault, sequela |
| T40.692A | Poisoning by other narcotics, intentional self-harm, initial encounter |
| T40.692D | Poisoning by other narcotics, intentional self-harm, subsequent encounter |
| T40.692S | Poisoning by other narcotics, intentional self-harm, sequela |
| T40.693A | Poisoning by other narcotics, assault, initial encounter |
| T40.693D | Poisoning by other narcotics, assault, subsequent encounter |
| T40.693S | Poisoning by other narcotics, assault, sequela |

Supplementary Table 2: UMLS Semantic Type categories used for CUI extraction.

| Abreviation | Code | Name |
|---|---|---|
| aggp | T100 | Age Group |
| antb | T195 | Antibiotic |

| | | |
|---|---|---|
| bacs | T123 | Biologically Active Substance |
| bhvr | T053 | Behavior |
| clna | T201 | Clinical Attribute |
| clnd | T200 | Clinical Drug |
| diap | T060 | Diagnostic Procedure |
| dsyn | T047 | Disease or Syndrome |
| hcro | T093 | Health Care Related Organization |
| hlca | T058 | Health Care Activity |
| lbpr | T059 | Laboratory Procedure |
| lbtr | T034 | Laboratory or Test Result |
| medd | T074 | Medical Device |
| neop | T191 | Neoplastic Process |
| ocac | T057 | Occupational Activity |
| ocdi | T090 | Occupation or Discipline |
| phsu | T121 | Pharmacologic Substance |
| socb | T054 | Social Behavior |
| sosy | T184 | Sign or Symptom |
| topp | T061 | Therapeutic or Preventive Procedure |

Supplementary Table 3: Demographic characteristics of the cohort of patients with at least one adverse opioid event relative to an average ED patient population. The opioid cohort was older with a mean age of 48.7 vs 42.0 ($p < 0.001$) for the non-opioid cohort. More predominantly male, 53.8% vs 45.9% ($p < 0.001$), and more predominantly Caucasian 65.0% vs 53.4% ($p < 0.001$).

| | | Overall | No Opioid Event | Opioid Exposure | P-Value |
|---|---|---|---|---|---|
| n | | 169207 | 86630 | 82577 | |
| Age(years), mean (SD) | | 45.3 (21.4) | 42.0 (25.4) | 48.7 (15.6) | <0.001 |
| Sex, n (%) | Female | 84987 (50.2) | 46884 (54.1) | 38103 (46.1) | <0.001 |
| Sex, n (%) | Male | 84196 (49.8) | 39740 (45.9) | 44456 (53.8) | |
| Sex, n (%) | Unknown | 24 (0.0) | 6 (0.0) | 18 (0.0) | |
| Race, n (%) | White or Caucasian | 99924 (59.1) | 46252 (53.4) | 53672 (65.0) | <0.001 |
| Race, n (%) | Black or African American | 38002 (22.5) | 20952 (24.2) | 17050 (20.6) | |
| Race, n (%) | Other | 26850 (15.9) | 16041 (18.5) | 10809 (13.1) | |
| Race, n (%) | Asian | 1884 (1.1) | 1627 (1.9) | 257 (0.3) | |
| Race, n (%) | Unknown | 1598 (0.9) | 1255 (1.4) | 343 (0.4) | |
| Race, n (%) | American Indian or Alaska Native | 581 (0.3) | 286 (0.3) | 295 (0.4) | |
| Race, n (%) | Native Hawaiian or Other Pacific Islander | 368 (0.2) | 217 (0.3) | 151 (0.2) | |
| Ethnicity, n (%) | Non-Hispanic | 133288 (78.8) | 65506 (75.6) | 67782 (82.1) | <0.001 |
| Ethnicity, n (%) | Hispanic or Latino | 34912 (20.6) | 20433 (23.6) | 14479 (17.5) | |
| Ethnicity, n (%) | Unknown | 1007 (0.6) | 691 (0.8) | 316 (0.4) | |

Supplementary Fig1: LDA Coherence Values and topics for other models

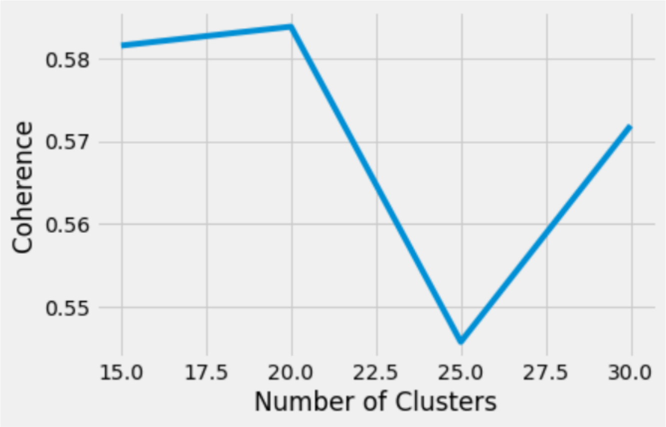


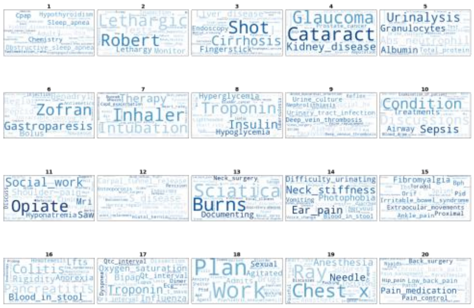

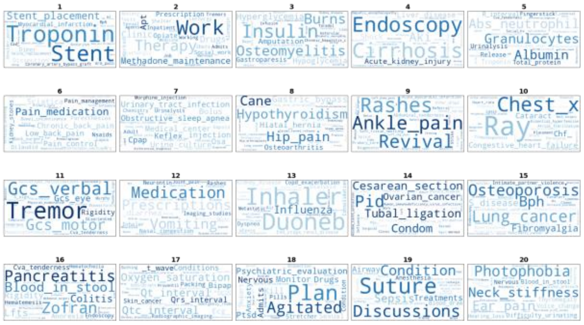

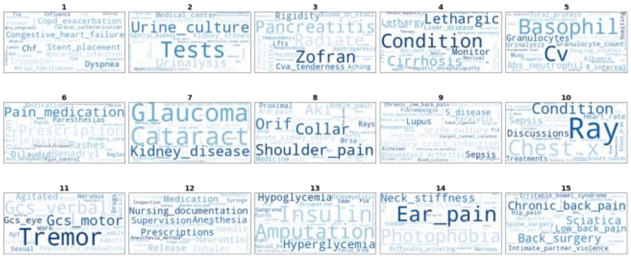

Supplementary Figure 2: Kmeans Clustering Outcomes

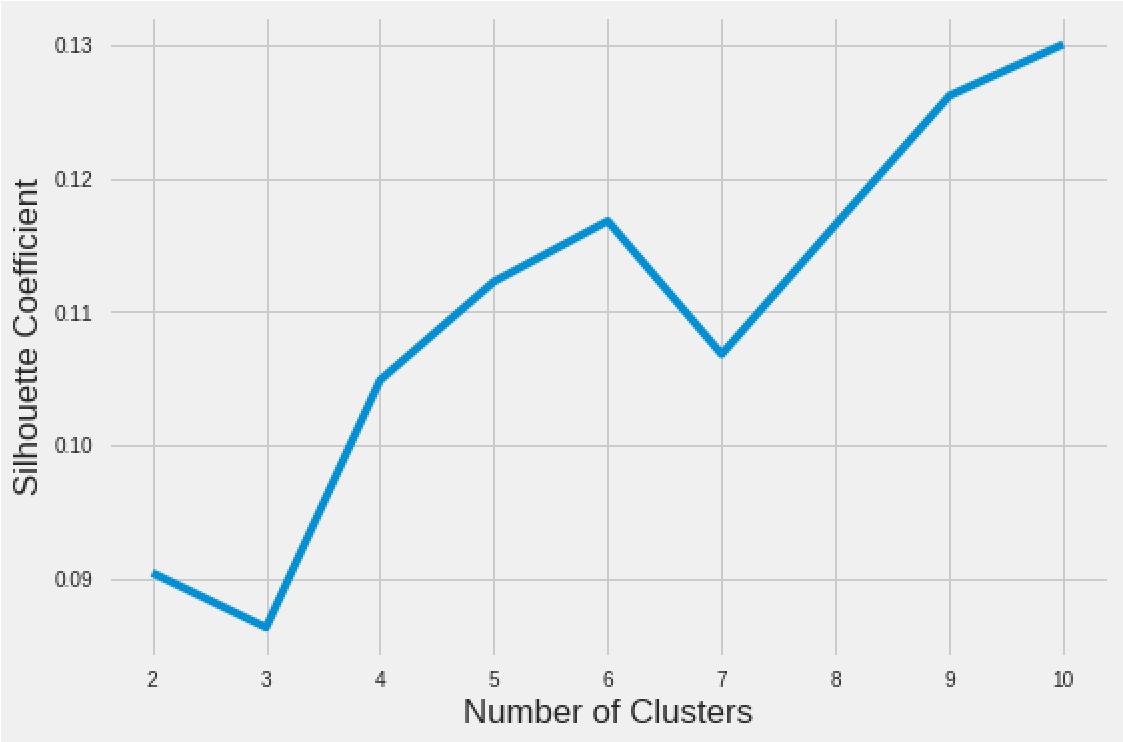


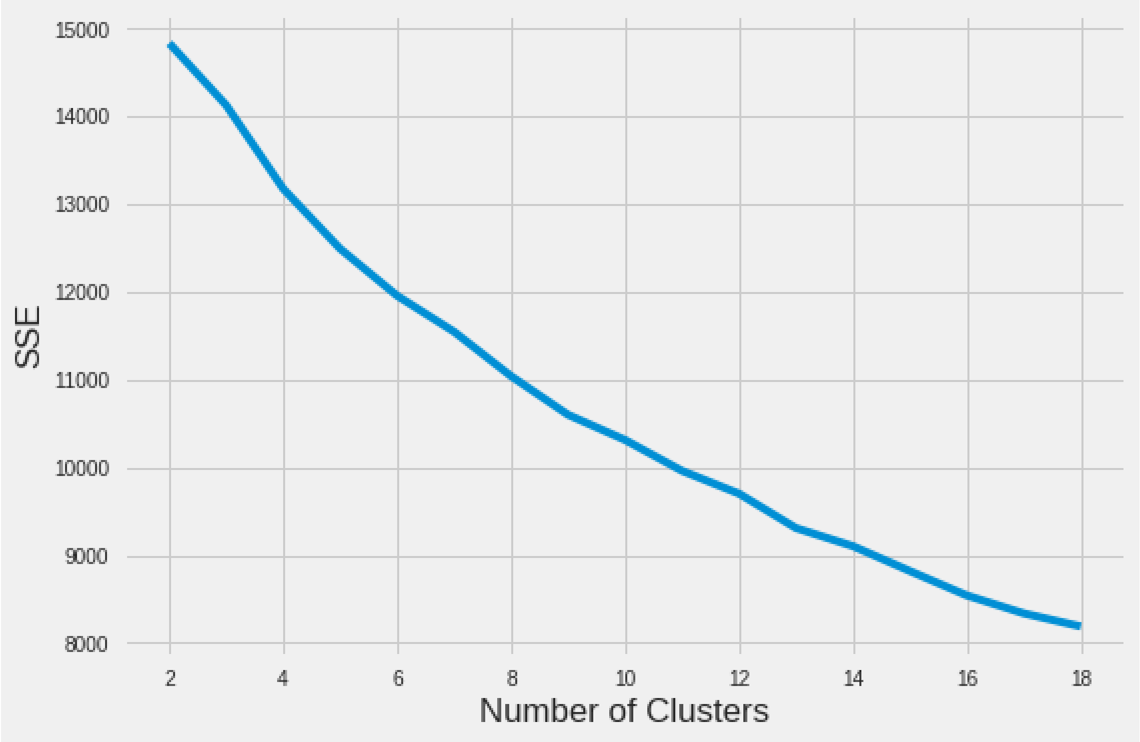
